# Risk of heart disease following breast cancer: results from a population-based cohort study

**DOI:** 10.1101/2021.09.16.21263682

**Authors:** Haomin Yang, Nirmala Bhoo-Pathy, Judith S. Brand, Elham Hedayati, Felix Grassmann, Erwei Zeng, Jonas Bergh, Weiwei Bian, Jonas F. Ludvigsson, Per Hall, Kamila Czene

## Abstract

**BACKGROUND:** There is a rising concern about treatment-associated cardiotoxicities in breast cancer patients.

**OBJECTIVES:** This study aimed to determine the time- and treatment-specific incidence of arrhythmia, heart failure and ischemic heart disease in women diagnosed with breast cancer.

**METHODS:** A register-based matched cohort study was conducted including 8338 breast cancer patients diagnosed from 2001-2008 in the Stockholm-Gotland region and followed-up until 2017. Overall and time dependent risks of arrhythmia, ischemic heart disease and heart failure in breast cancer patients were assessed using flexible parametric models as compared to matched controls from general population. Treatment-specific effects were estimated in breast cancer patients using Cox model.

**RESULTS:** During a median follow-up of 10.8 years, the hazard ratios for arrhythmia, heart failure and ischemic heart disease, were 1.27 (95% CI = 1.18-1.37), 1.38 (95% CI = 1.23-1.55), and 0.93 (95% CI = 0.84-1.03), respectively. Time-dependent analyses revealed long-term increased risks of arrhythmia, and heart failure in breast cancer patients compared to matched controls. The risk of ischemic heart disease was only elevated in the first year after cancer diagnosis. Trastuzumab and anthracyclines were associated with increased risk of heart failure. Aromatase inhibitors, but not tamoxifen, were associated with risk of ischemic heart disease. No increased risk of heart disease was identified following loco-regional radiotherapy.

**CONCLUSIONS:** Administration of systemic adjuvant therapies appear to be associated with increased risks of heart disease. The risk estimates observed in this study may serve as reference to aid adjuvant therapy decision-making and patient counseling in oncology practices.

## INTRODUCTION

The use of adjuvant systemic therapies at least halves the risk of dying from breast cancer (Early Breast Cancer Trialists’ Collaborative, 2005, 2012; Early Breast Cancer Trialists’ Collaborative et al., 2015; Goldvaser et al., 2019; Gray et al., 2019). Nowadays, 80% of the breast cancer patients survive for at least 10 years and many will become long-term survivors (Colzani et al., 2011a). There are, however, concerns about therapy associated late adverse health effects, including cardiovascular events (Khouri et al., 2012). Use of common (neo-) adjuvant therapies for breast cancer have been associated with an increased risk of heart diseases including heart failure, arrhythmias and ischemic heart disease (Darby et al., 2013; Doyle, Neugut, Jacobson, Grann, & Hershman, 2005; Harris et al., 2006; Hooning et al., 2007; Pinder, Duan, Goodwin, Hortobagyi, & Giordano, 2007; C. Taylor et al., 2017; Yeh & Bickford, 2009). However, this evidence mostly comes from studies focusing on specific subgroups of patients based on age, cancer stage or treatment regimen.

Although the benefits of radiotherapy far outweigh the risk of heart diseases, some studies have found increased incidence of and mortality due to heart disease in women subjected to modern doses of adjuvant radiotherapy (Darby et al., 2013; C. Taylor et al., 2017). Knowledge on cardiotoxic effects of anthracycline-based chemotherapy regimens have led to lowering of doses and less use of bolus injections to reduce peak concentrations of anthracyclines (Foukakis et al., 2016), but it is estimated that the risk of heart failure associated with anthracyclines remains increased for standard low-dose group as compared to non-users (Chung et al., 2020). While trastuzumab has been shown to reduce the risk of breast cancer mortality at 11 years of follow-up (Cameron et al., 2017), evidence on its cardiotoxicity is conflicting (Bowles et al., 2012; Papakonstantinou et al., 2020). In addition, recent evidence suggests that use of aromatase inhibitors in women with hormone receptor positive breast cancer may increase the risk of heart failure, compared to tamoxifen (Khosrow-Khavar, Filion, Bouganim, Suissa, & Azoulay, 2020).

Risk assessment of immediate and later occurring heart disease events following breast cancer is important for the planning of cardiac surveillance programs and possible prophylactic pharmacotherapy. Here, we report the risks of heart diseases in in a cohort representative of the general breast cancer population with long-term follow-up. We specifically aimed to assess risks of heart diseases by time since diagnosis, and according to adjuvant treatments.

## Materials and Methods

### Breast cancer cohort

This study took advantage of the Stockholm-Gotland Breast Cancer Register comprising all women diagnosed with primary invasive breast cancer between 2001 and 2008 in the Stockholm-Gotland region. The Stockholm-Gotland Breast Cancer Register has about 99% completeness and provides detailed information on tumor and treatment characteristics, as well as routine follow-up on locoregional recurrences and distant metastases (Colzani et al., 2011b; Wigertz et al., 2012). We included all patients diagnosed with non-metastatic disease (stages I-III) at age 25 to 75 years (N = 8338). To compare the risk of heart diseases after breast cancer, we randomly sampled up to 10 women from the general female population in Stockholm-Gotland region matched on year of birth and whether they had history of the three heart diseases (Supplementary Figure 1). We performed the matching procedure for these three heart diseases separately. Each reference individual was alive and free of breast cancer on the date of the matched patient’s diagnosis of breast cancer (the index date).

The matched cohort was linked through the unique personal identity number to the Swedish Cancer Register, Patient Register, Cause of Death Register and Migration Register, and follow-up started from the index date until the date of heart disease diagnosis, emigration, death, breast cancer relapse or end of follow-up (31 December 2017), whichever occurred first. We also performed linkage with the Prescribed Drug Register which contains data on all drugs dispensed from Swedish pharmacies from July 2005 onward to validate the use of hormone therapy. Detailed description of the breast cancer cohort can be found elsewhere (Holm et al., 2016).

The study was approved by the Regional Ethical Review Board in Stockholm.

### Heart diseases

We identified the following heart diseases according to relevant ICD (International Classification of Disease) codes in the Swedish Patient Register and the Cause of Death Register (Supplementary Table 1): heart failure (ICD-10: I50, ICD-9: 428A, 428B, 428X), arrhythmias (ICD-10: I47-I49, ICD-9: 427), and ischemic heart disease (ICD-10: I20-I25, ICD-9: 410-414). We included both inpatient and outpatient diagnoses, as well as cause of death records in our outcome definition, except for myocardial infarction, which was based on inpatient and cause of death records solely. To ensure specificity of the outcomes studied, only primary, and not underlying, diagnoses were considered for analyses.

### Breast cancer treatment specifics

We extracted the treatment data from the Stockholm-Gotland Breast Cancer Register. Because ∼90% of HER-2 positive cancers were treated with trastuzumab between 2005 and 2008 in the Stockholm-Gotland region, HER-2 positivity was used as a proxy when no registry data on trastuzumab was available during this time period. Data on adjuvant endocrine therapy was verified against the Prescribed Drug Register and categorized into tamoxifen and / or aromatase inhibitor (AI) use. Since radiotherapy to the left breast has, in particular, been implicated in heart complications, radiotherapy was categorized according to tumor laterality (left vs. right). Bilateral tumors were coded as receiving left-sided radiotherapy in this analysis. Chemotherapy was coded as anthracycline-based, anthracycline plus taxane-based, cyclophosphamide, methotrexate and fluorouracil (CMF)-based regimens, or unknown/unspecified.

### Covariates

The Stockholm Breast Cancer Register contains data on date of diagnosis, menopausal status at diagnosis and type of surgery (breast conserving surgery vs. mastectomy). Tumor characteristics were also retrieved from this register, including tumor size (T), regional lymph node involvement (N) and presence of metastases (M), all from pathology records and summarized in TNM stage as defined according to the American Joint Committee on Cancer (“American Joint Committee on Cancer. Breast Cancer Staging. 7th Edition.,”). Information on inpatient comorbidities at diagnosis was extracted though the Swedish Patient Register and summarized into the Charlson Comorbidity Index (CCI) score, a widely used method for classifying chronic comorbid conditions (Charlson, Pompei, Ales, & MacKenzie, 1987). To account for the potential confounding effect from tobacco abuse, chronic pulmonary disease, and hypertension in the associations, we further identified associated diagnoses before cancer using ICD codes from the patient register. (Supplementary Table 1)

### Statistical analyses

We compared the risk of heart diseases in breast cancer patients with that observed in the matched cohort, using flexible parametric model (FPM), overall and stratified by history of respective heart disease and time since diagnosis. We further plotted time-dependent hazard ratios using FPM. The FPM is similar to the Cox proportional hazards model in that it provides a HR as measure of association. The key advantage of FPM is that non-proportional hazards can easily be fitted by adding a spline for the interaction with time. Kaplan-Meier curves were used to assess the cumulative incidences of heart diseases in breast cancer patients and matched reference individuals.

Next, we studied the association of adjuvant breast cancer therapy with heart disease risk in breast cancer patients using Cox proportional hazards models. We adjusted these analyses for age and year of diagnosis (model 1), and additionally for menopausal status at diagnosis, cancer stage, type of surgery, CCI score, history of heart disease, hypertension, chronic pulmonary disease and tobacco abuse. All treatment specific models were mutually adjusted for adjuvant therapies. Taking the possible selection bias in administration of radiotherapy into consideration, the analysis for radiotherapy only included patients receiving radiotherapy, making a comparison between left-sided and right-sided breast cancer (Wadsten et al., 2018). Missing indicators were included for the analysis of these covariates in the model. To investigate the robustness of treatment specific associations, a sensitivity analysis was performed excluding breast cancer patients with a history of heart disease. To explore the potential time dependent effect of treatment, we separated the analysis according to different follow-up periods, within 10 years after breast cancer diagnosis and beyond, respectively.

All statistical analyses were performed using STATA version 15.1.

## RESULTS

Descriptive characteristics of the study population are described in Table 1. The median age at breast cancer diagnosis was 59 years, with 75% of patients aged less than 65 years (Table 1). Approximately 40% of all patients received adjuvant chemotherapy, with anthracyline-based regimens being most frequently administered. Endocrine therapy was received by more than 80% of patients (constituting mainly of tamoxifen). Of all patients diagnosed between mid-2005 and 2008, 13% were coded as receivers of trastuzumab.

**Table 1.**
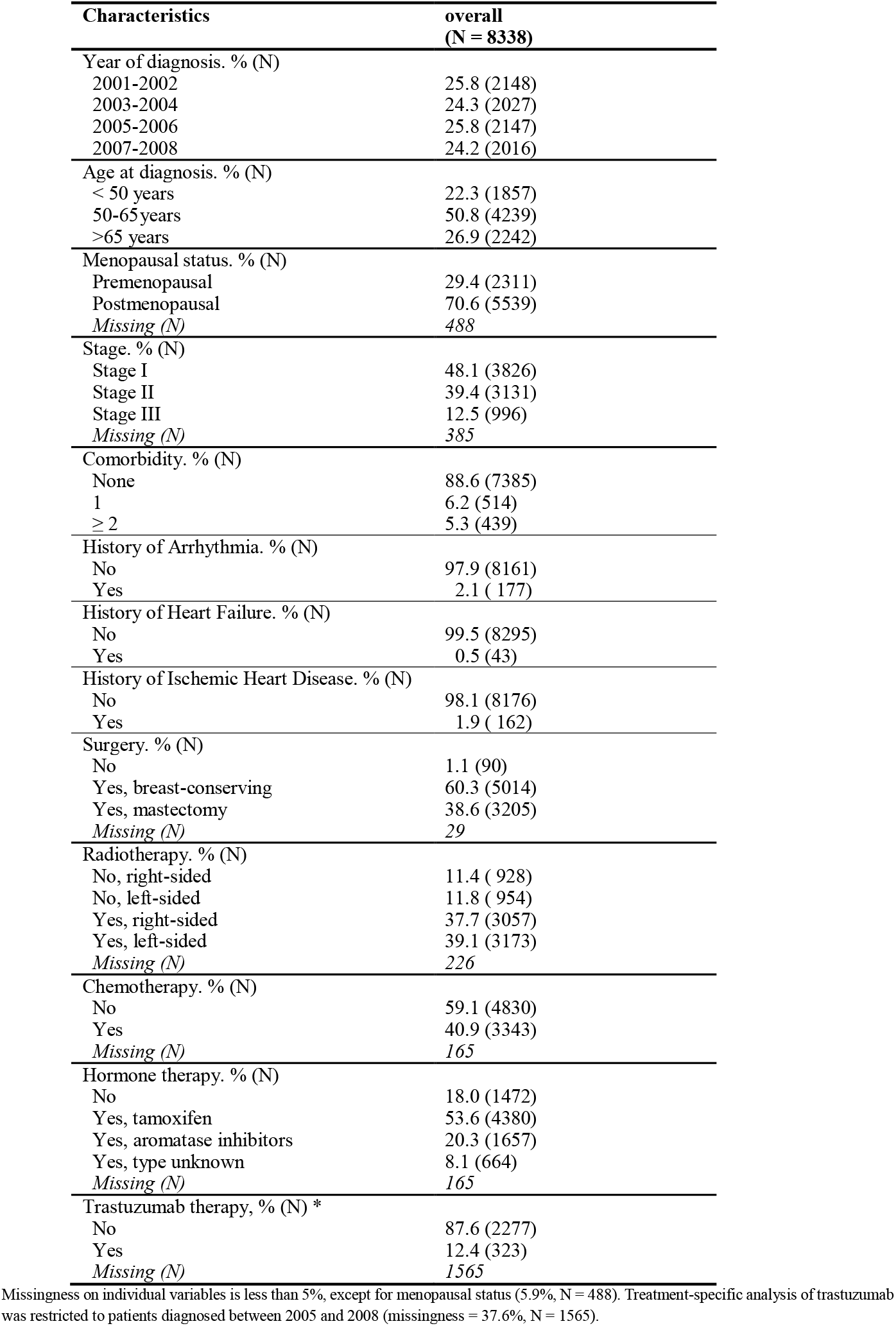
Descriptive characteristics of the study population.

| **Characteristics** | **overall**<br>**(N = 8338)** |
| --- | --- |
| Year of diagnosis. % (N) |  |
| 2001-2002 | 25.8 (2148) |
| 2003-2004 | 24.3 (2027) |
| 2005-2006 | 25.8 (2147) |
| 2007-2008 | 24.2 (2016) |
| Age at diagnosis. % (N) |  |
| < 50 years | 22.3 (1857) |
| 50-65 years | 50.8 (4239) |
| >65 years | 26.9 (2242) |
| Menopausal status. % (N) |  |
| Premenopausal | 29.4 (2311) |
| Postmenopausal | 70.6 (5539) |
| Missing (N) | 488 |
| Stage. % (N) |  |
| Stage I | 48.1 (3826) |
| Stage II | 39.4 (3131) |
| Stage III | 12.5 (996) |
| Missing (N) | 385 |
| Comorbidity. % (N) |  |
| None | 88.6 (7385) |
| 1 | 6.2 (514) |
| ≥ 2 | 5.3 (439) |
| History of Arrhythmia. % (N) |  |
| No | 97.9 (8161) |
| Yes | 2.1 ( 177) |
| History of Heart Failure. % (N) |  |
| No | 99.5 (8295) |
| Yes | 0.5 (43) |
| History of Ischemic Heart Disease. % (N) |  |
| No | 98.1 (8176) |
| Yes | 1.9 ( 162) |
| Surgery. % (N) |  |
| No | 1.1 (90) |
| Yes, breast-conserving | 60.3 (5014) |
| Yes, mastectomy | 38.6 (3205) |
| Missing (N) | 29 |
| Radiotherapy. % (N) |  |
| No, right-sided | 11.4 ( 928) |
| No, left-sided | 11.8 ( 954) |
| Yes, right-sided | 37.7 (3057) |
| Yes, left-sided | 39.1 (3173) |
| Missing (N) | 226 |
| Chemotherapy. % (N) |  |
| No | 59.1 (4830) |
| Yes | 40.9 (3343) |
| Missing (N) | 165 |
| Hormone therapy. % (N) |  |
| No | 18.0 (1472) |
| Yes, tamoxifen | 53.6 (4380) |
| Yes, aromatase inhibitors | 20.3 (1657) |
| Yes, type unknown | 8.1 (664) |
| Missing (N) | 165 |
| Trastuzumab therapy, % (N) * |  |
| No | 87.6 (2277) |
| Yes | 12.4 (323) |
| Missing (N) | 1565 |

Missingness on individual variables is less than 5%, except for menopausal status (5.9%, N = 488). Treatment-specific analysis of trastuzumab was restricted to patients diagnosed between 2005 and 2008 (missingness = 37.6%, N = 1565).

Over a median follow-up of 10.8 years (interquartile range = 6.5 years) arrhythmias were the most frequently reported heart disease (n = 677), followed by ischemic heart disease (n = 384) and heart failure (n = 306). The cumulative incidence of arrhythmias, ischemic heart disease and heart failure in the cohort was 12.10%, 6.55% and 5.59% after 15 years of follow-up, respectively (Supplementary Figure 2 and Supplementary Table 2).

Overall, breast cancer patients experienced a 27% increased risk (HR = 1.27; 95% CI = 1.18-1.37) of developing arrhythmia, and a 38% increased risk of heart failure (HR=1.38; 95% CI = 1.23-1.55) compared to matched reference individuals from general population, while the risk of ischemic heart disease was not increased (HR = 0.93; 95% CI = 0.84-1.03) (Table 2). Hazard ratios of for arrhythmias, ischemic heart disease and heart failure were not notably different in women with or without a previous diagnosis of these respective heart diseases.

**Table 2.** Hazard for heart diseases in breast cancer patients compared to the matched cohort.

|  | Arrhythmia |  | Heart Failure |  | Ischemic Heart Disease |  |
| --- | --- | --- | --- | --- | --- | --- |
|  | No. | HR (95% CI) | No. | HR (95% CI) | No. | HR (95% CI) |
| **Overall** | 677 | **1.27 (1.18-1.37)** | 306 | **1.38 (1.23-1.55)** | 384 | 0.93 (0.84-1.03) |
| **Corresponding disease history** |  |  |  |  |  |  |
| **No** | 585 | **1.28 (1.18-1.39)** | 286 | **1.39 (1.23-1.57)** | 321 | 0.91 (0.81-1.01) |
| **Yes** | 92 | **1.30 (1.04-1.61)** | 20 | 1.25 (0.77-2.05) | 63 | 1.04 (0.80-1.36) |
| **Time since diagnosis** |  |  |  |  |  |  |
| **<1 year** | 109 | **2.21 (1.81-2.70)** | **37** | **2.71 (1.88-3.91)** | **65** | **1.48 (1.14-1.91)** |
| **1-2 years** | 48 | 1.10 (0.82-1.48) | 24 | **1.94 (1.26-3.01)** | 41 | 0.97 (0.70-1.34) |
| **2-5 years** | 123 | 1.03 (0.86-1.24) | 52 | 1.31 (0.99-1.74) | 94 | 0.92 (0.75-1.13) |
| **5-10 years** | 226 | 1.13 (0.99-1.30) | 95 | 1.13 (0.91-1.38) | 121 | 0.84 (0.70-1.00) |
| **10-17 years** | 171 | **1.42 (1.22-1.66)** | 98 | **1.38 (1.12-1.69)** | 63 | 0.79 (0.61-1.01) |

\* Abbreviations: No.=number of cases. HR=hazard ratio. CI = confidence interval. The HRs are estimated using flexible parametric model and conditioned on matching criteria (year of birth and history of respective heart disease). In all models, time since index date was the underlying time scale and a restricted cubic spline with four internal and two boundary knots (five degrees of freedom) placed at quintiles of the event times, was used for the baseline hazard.

Analyses by time since diagnosis revealed a short-term increase in risks of arrhythmia and heart failure (Table 2). An elevated risk of ischemic heart disease was observed in the first year after diagnosis (HR = 1.48; 95% CI = 1.14-1.91), followed by a decline with increasing follow-up time. Interestingly, increased risks for arrhythmia and heart failure were noted beyond 10 years (Table 2, Figure 1).

**Figure 1.**
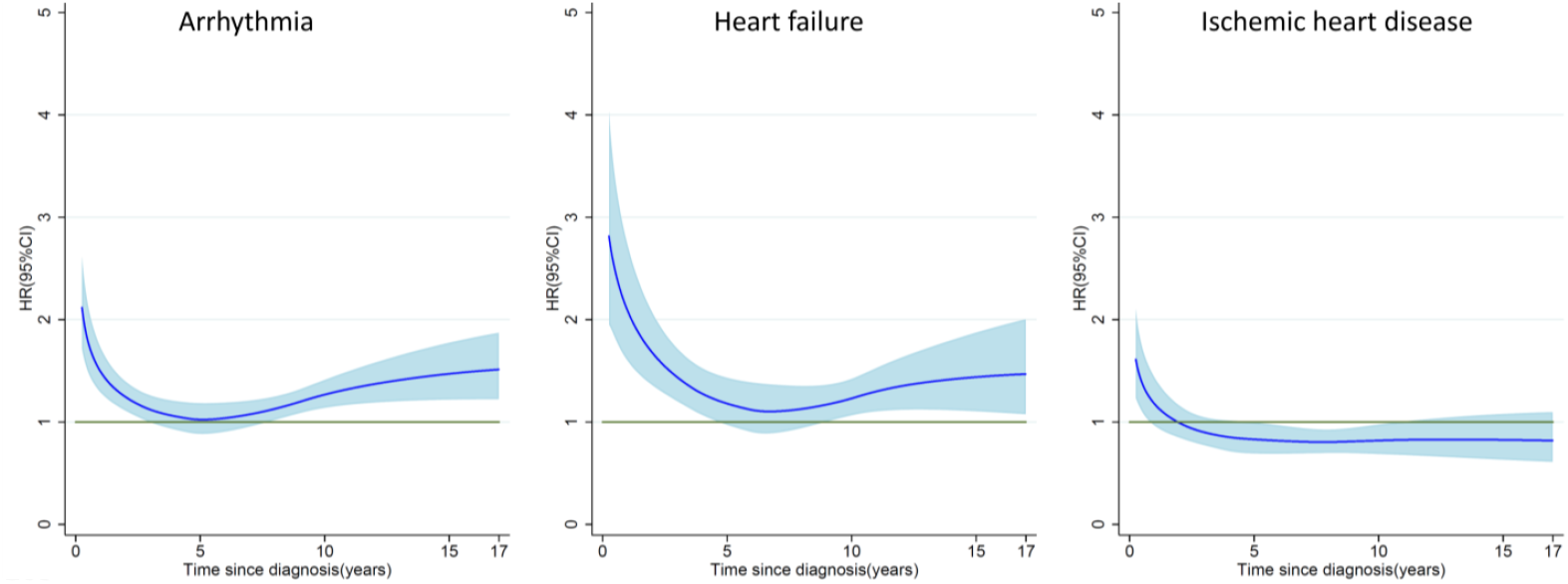
Time dependent hazard ratio of heart diseases in breast cancer patients comparing to age-matched women. In all models, time since index date was the underlying time scale and a restricted cubic spline with four internal and two boundary knots (five degrees of freedom) placed at quintiles of the event times, was used for the baseline hazard. Time-dependent effects were modelled by adding interaction terms with time using a second spline with three degrees of freedom.

Patients treated with chemotherapy and trastuzumab had an increased risk of heart failure compared to patients not receiving these treatments (for patients receiving anthracyclines-based chemotherapy, HR=1.99, 95% CI=1.38-2.85; for patients receiving trastuzumab, HR=2.04, 95%CI=1.10-3.80) (Table 3). The risk of heart failure in patients receiving anthracyclines ever persisted for 10 years beyond the diagnosis of breast cancer (HR= 2.31, 95%CI=1.21-4.43, supplementary table 3). Receipt of aromatase inhibitors was marginally associated with risk of ischemic heart disease (HR=1.42, 95%CI=1.00-2.01 compared to patients without hormonal therapy), but not with risk of heart failure or arrhythmias (Table 3). A direct comparison between left-sided and right-sided radiotherapy, showed no evidence of an association of radiotherapy with heart disease except for a somewhat increased risk of ischemic heart disease in women with left sided breast cancer, particularly after 10 years after cancer diagnosis, although not statistically significant (HR= 1.14, 95%CI=0.89-1.46; HR for the risk beyond 10 years=1.32, 95%CI=0.74-2.36). Sensitivity analyses excluding breast cancer patients with a previous history of heart disease only slightly changed the point estimates of the treatment effects but not the inferences (Supplementary Table 4).

**Table 3.**
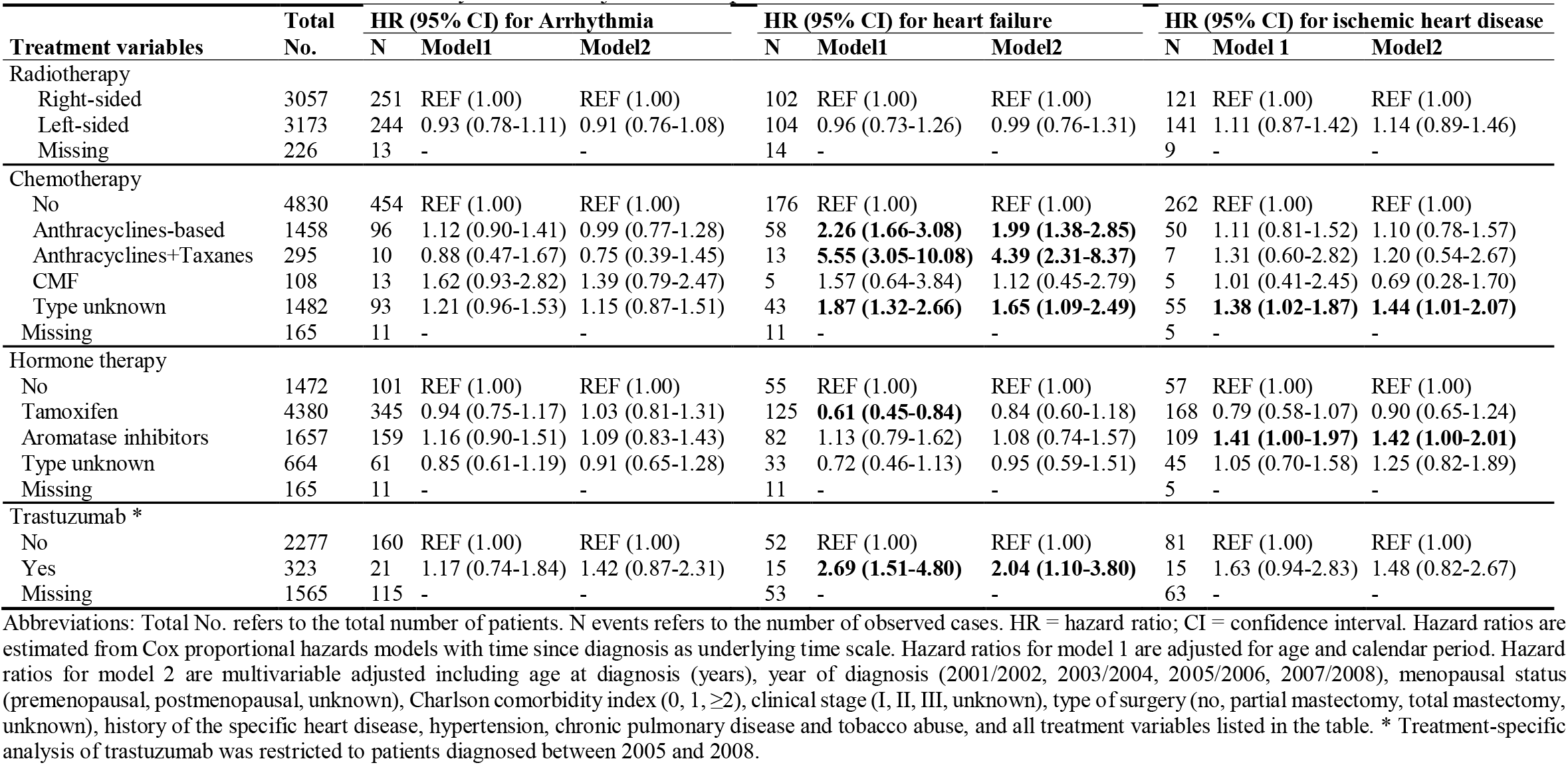
Hazard ratios for heart diseases by different adjuvant therapies

| Treatment variables | Total No. | HR (95% CI) for Arrhythmia |  |  | HR (95% CI) for heart failure |  |  | HR (95% CI) for ischemic heart disease |  |  |
| --- | --- | --- | --- | --- | --- | --- | --- | --- | --- | --- |
|  |  | N | Model1 | Model2 | N | Model1 | Model2 | N | Model 1 | Model2 |
| Radiotherapy |  |  |  |  |  |  |  |  |  |  |
| Right-sided | 3057 | 251 | REF (1.00) | REF (1.00) | 102 | REF (1.00) | REF (1.00) | 121 | REF (1.00) | REF (1.00) |
| Left-sided | 3173 | 244 | 0.93 (0.78-1.11) | 0.91 (0.76-1.08) | 104 | 0.96 (0.73-1.26) | 0.99 (0.76-1.31) | 141 | 1.11 (0.87-1.42) | 1.14 (0.89-1.46) |
| Missing | 226 | 13 | - | - | 14 | - | - | 9 | - | - |
| Chemotherapy |  |  |  |  |  |  |  |  |  |  |
| No | 4830 | 454 | REF (1.00) | REF (1.00) | 176 | REF (1.00) | REF (1.00) | 262 | REF (1.00) | REF (1.00) |
| Anthracyclines-based | 1458 | 96 | 1.12 (0.90-1.41) | 0.99 (0.77-1.28) | 58 | **2.26 (1.66-3.08)** | **1.99 (1.38-2.85)** | 50 | 1.11 (0.81-1.52) | 1.10 (0.78-1.57) |
| Anthracyclines+Taxanes | 295 | 10 | 0.88 (0.47-1.67) | 0.75 (0.39-1.45) | 13 | **5.55 (3.05-10.08)** | **4.39 (2.31-8.37)** | 7 | 1.31 (0.60-2.82) | 1.20 (0.54-2.67) |
| CMF | 108 | 13 | 1.62 (0.93-2.82) | 1.39 (0.79-2.47) | 5 | 1.57 (0.64-3.84) | 1.12 (0.45-2.79) | 5 | 1.01 (0.41-2.45) | 0.69 (0.28-1.70) |
| Type unknown | 1482 | 93 | 1.21 (0.96-1.53) | 1.15 (0.87-1.51) | 43 | **1.87 (1.32-2.66)** | **1.65 (1.09-2.49)** | 55 | **1.38 (1.02-1.87)** | **1.44 (1.01-2.07)** |
| Missing | 165 | 11 | - | - | 11 | - | - | 5 | - | - |
| Hormone therapy |  |  |  |  |  |  |  |  |  |  |
| No | 1472 | 101 | REF (1.00) | REF (1.00) | 55 | REF (1.00) | REF (1.00) | 57 | REF (1.00) | REF (1.00) |
| Tamoxifen | 4380 | 345 | 0.94 (0.75-1.17) | 1.03 (0.81-1.31) | 125 | **0.61 (0.45-0.84)** | 0.84 (0.60-1.18) | 168 | 0.79 (0.58-1.07) | 0.90 (0.65-1.24) |
| Aromatase inhibitors | 1657 | 159 | 1.16 (0.90-1.51) | 1.09 (0.83-1.43) | 82 | 1.13 (0.79-1.62) | 1.08 (0.74-1.57) | 109 | **1.41 (1.00-1.97)** | **1.42 (1.00-2.01)** |
| Type unknown | 664 | 61 | 0.85 (0.61-1.19) | 0.91 (0.65-1.28) | 33 | 0.72 (0.46-1.13) | 0.95 (0.59-1.51) | 45 | 1.05 (0.70-1.58) | 1.25 (0.82-1.89) |
| Missing | 165 | 11 | - | - | 11 | - | - | 5 | - | - |
| Trastuzumab * |  |  |  |  |  |  |  |  |  |  |
| No | 2277 | 160 | REF (1.00) | REF (1.00) | 52 | REF (1.00) | REF (1.00) | 81 | REF (1.00) | REF (1.00) |
| Yes | 323 | 21 | 1.17 (0.74-1.84) | 1.42 (0.87-2.31) | 15 | **2.69 (1.51-4.80)** | **2.04 (1.10-3.80)** | 15 | 1.63 (0.94-2.83) | 1.48 (0.82-2.67) |
| Missing | 1565 | 115 | - | - | 53 | - | - | 63 | - | - |

Abbreviations: Total No. refers to the total number of patients. N events refers to the number of observed cases. HR = hazard ratio; CI = confidence interval. Hazard ratios are estimated from Cox proportional hazards models with time since diagnosis as underlying time scale. Hazard ratios for model 1 are adjusted for age and calendar period. Hazard ratios for model 2 are multivariable adjusted including age at diagnosis (years), year of diagnosis (2001/2002, 2003/2004, 2005/2006, 2007/2008), menopausal status (premenopausal, postmenopausal, unknown), Charlson comorbidity index (0, 1, $\geq 2$ ), clinical stage (I, II, III, unknown), type of surgery (no, partial mastectomy, total mastectomy, unknown), history of the specific heart disease, hypertension, chronic pulmonary disease and tobacco abuse, and all treatment variables listed in the table. \* Treatment-specific analysis of trastuzumab was restricted to patients diagnosed between 2005 and 2008.

## DISCUSSION

In a population-based setting, we demonstrated that the incidence of heart disease in breast cancer patients was significantly higher than the incidence observed in matched reference individuals from the general population. The risks of arrhythmia and heart failure were increased even beyond 15 years after diagnosis. Receipt of trastuzumab, as well as administration of anthracycline +/-taxane-based regimens were independently associated with an increased risk of heart failure, while aromatase inhibitor therapy was associated with an increased risk of ischemic heart disease.

We found an increased risk of arrhythmia and heart failure in breast cancer patients as compared with the matched reference individuals from the general population, which is similar to the risk of heart failure reported by a previous Dutch study (Hooning et al., 2007), indicating the generalizability of our findings within European countries. In the present study, we further found similar magnitude HRs estimates in patients with and without a previous diagnosis of heart disease, suggesting no effect modification by disease history.

Analysis by time since diagnosis from the current study revealed long-term increased risks of arrhythmia and heart failure following breast cancer. The long-term increased risk of heart failure potentially reflects the profound cardiotoxic effect of anthracyclines, as we found increased risk of heart failure in patients treated with anthracyclines beyond 10 years after cancer diagnosis. As the long-term risk was observed for heart failure but not ischemic heart disease, we hypothesized that the cardiotoxic effect of chemotherapy was mainly on the myocardium but not the cardiac vessels. The finding that risk of ischemic heart disease in breast cancer patients may be shortly elevated after diagnosis, but diminish over time is not unexpected, considering the emotional distress of dealing with a new cancer diagnosis in the patients, which may lead to higher short-term rates of ischemic heart disease (Fang et al., 2012; Schoormans, Pedersen, Dalton, Rottmann, & van de Poll-Franse, 2016).

The findings that administration of anthracycline +/-taxanes, as well as trastuzumab are associated with increased risks of heart failure are in agreement with previous observational studies (Bowles et al., 2012; Chavez-MacGregor et al., 2013; Doyle et al., 2005; Du et al., 2009; Goldhar et al., 2016; Thavendiranathan et al., 2016) and clinical trials (Chen et al., 2011) in specific subgroups of breast cancer patients. While (poly)chemotherapy is thought to lead to heart failure through a plethora of mechanisms that damage cardiomyocytes (Yeh & Bickford, 2009), taxanes may also interrupt the metabolism and excretion of anthracyclines, hence accentuating their toxicity (Bird & Swain, 2008). Inhibition of the HER2 receptors in myocytes, which regulates cell development and growth may largely explain the cardiotoxicity caused by trastuzumab (Yeh & Bickford, 2009).

Population-based estimates of incidence of ischemic heart disease following treatment with aromatase inhibitors have not been extensively reported in the literature (Matthews et al., 2018). Abdel-Qadir, et al also found an increased risk of myocardial infarction in patients treated with aromatase inhibitors, as compared to those treated with tamoxifen (Abdel-Qadir et al., 2016). It has been posited that this apparent increase in risk of ischemic heart disease might in fact be attributed to reduced risk of heart disease following tamoxifen treatment (Hackshaw et al., 2011; Rosell et al., 2013). Nonetheless, given that in the present study, an increased risk of ischemic heart disease was observed in patients treated with aromatase inhibitors compared to patients not receiving hormone treatment, prior hypothesis that this observation is explained by the protective effect of tamoxifen is not fully correct.

While several previous studies have shown that radiotherapy for breast cancer increase the risk of ischemic heart disease and heart failure (Darby et al., 2013; Harris et al., 2006; Pinder et al., 2007), some studies, particularly those including patients diagnosed in recent decades, have not seen such increase (Boekel et al., 2014; Wadsten et al., 2018; Wennstig et al., 2020). Indeed, the time trends of heart dose improvement and the use of modern heart dose sparing techniques, together with individualized doses of therapy, may result in lower doses of radiation to the heart (C. Taylor et al., 2017; C. W. Taylor & Kirby, 2015; C. W. Taylor et al., 2009). Another reason for inconsistent results in the literature could be that patients with a left sided breast cancer who are susceptible to cardiovascular disease are less likely to be selected for radiotherapy (C. W. Taylor & Kirby, 2015). Therefore, in our study, we compared risk of ischemic heart disease in patients treated with radiotherapy for left-sided to those treated for right-sided breast cancer. We found slightly increased non-significant risk of ischemic heart disease, this risk appeared (as expected) more pronounced in the follow-up period after 10 years. Overall, our results indicate only small risk of heart disease due to radiotherapy in women treated in Sweden after year 2000.

A major strength of our study is that we were able to investigate risks of heart disease by time since diagnosis and adjuvant treatment. The risk estimates presented in this study therefore provide an insight into the short-term and long-term unintended effects of adjuvant breast cancer therapy in routine clinical practice. Notably, all breast cancer patients were followed-up for a relatively long period. The population-based nature of the study and linkage to registry-based data further minimized the risk of selection and information biases.

We acknowledge several limitations. The Swedish Patient Register has high validity for heart failure, arrhythmia and ischemic heart disease (with positive predictive value between 88%-98%) (Hammar et al., 2001; Ludvigsson et al., 2011), but some misclassification of heart diseases may have occurred. We further tried to reduce the likelihood of misclassification by analysing main diagnoses only. Secondly, we did not have complete data on trastuzumab during the study period and relied on HER-2 status as proxy, which may have resulted in some misclassification. The numbers of heart disease after trastuzumab and taxane were also relatively small in our study, and might have resulted in limited power or chance finding when identifying an association between trastuzumab/taxane and the relevant cardiac events. Besides, the Stockholm-Gotland Breast Cancer Register only records intended treatment with radiotherapy and chemotherapy, not whether patients were actually administrated with these therapies. Finally, it should be noted that cancer patients are subject to increased medical surveillance, especially in the initial period after diagnosis. This could explain the increased rates of heart disease events especially within the first year of follow-up, although psychological distress following recent diagnosis with cancer represents an alternative plausible explanation for this observation (Fang et al., 2012; Schoormans et al., 2016).

Through this study, we demonstrate that compared to the general population, women with breast cancer are associated with significantly increased risks of cardiovascular events including heart failure and arrhythmia. The short term risk of ischemic heart disease diminished after 1 year post diagnosis. The increased risk of arrhythmia and heart failure however appears to persist long-term, beyond the first decade after diagnosis. The risk estimates observed in this study may serve as reference to aid adjuvant therapy decision-making and patient counselling in oncology practices.

## Data Availability

Data Availability: The data used in this study are owned by the Swedish National Board of Health and Welfare and Statistics Sweden. According to Swedish law and GDPR, the authors are not able to make the dataset publicly available. Any researchers (including international researchers) interested in obtaining the data can do so by the following steps: 1) apply for ethical approval from their local ethical review boards; 2) contact the Swedish National Board of Health and Welfare and/or Statistics Sweden with the ethical approval and make a formal application of use of register data.

## LIST OF ABBREVIATIONS

ICD: International Classification of Diseases
CI: confidence intervals
HR: hazard ratio
CCI: Charlson Comorbidity Index

## DECLARATIONS

### Ethics approval and consent to participate

The study was approved by the Regional Ethical Review Board in Stockholm (Dnr 2009/254-31/4).

### Author contributions

HY had full access to all of the data in the study and takes responsibility for the integrity of the data and the accuracy of the data analysis. KC, NBP and HY conceived and designed the study. All authors acquired, analyzed, or interpreted the data. HY and NBP drafted the manuscript. All authors critically revised the manuscript for important intellectual content. HY performed the statistical analysis. KC, NBP and HY obtained the funding. All authors read and approved the final manuscript.

## Acknowledgements

Not applicable.

## Availability of data and materials

The register-based cohort datasets linked and analyzed in the current study are not publicly available due to Swedish law, but are available by applying through the Swedish National Board of Health and Welfare and Statistics Sweden. Detailed information on data application can be found using the following links https://bestalladata.socialstyrelsen.se/data-for-forskning/ and https://www.scb.se/vara-tjanster/bestalla-mikrodata/.

## Consent to publish

All authors approved the manuscript and consented to its publication.

## Authors’ information

Not applicable.

## ONLINE SUPPLEMENTARY

**Supplementary Table 1**. ICD codes used in the analyses

**Supplementary Table 2**. Cumulative incidence estimates of heart diseases in breast cancer patients and age matched controls.

**Supplementary Table 3**. Hazard ratios for heart diseases by different adjuvant therapies and time since diagnosis.

**Supplementary Table 4**. Hazard ratios for heart diseases by different adjuvant therapies, excluding patients with a history of heart diseases.

**Supplementary Figure 1**. Flow chart of the data.

**Supplementary Figure 2**. Cumulative incidence of heart disease in breast cancer patients and matched women.

Kaplan-Meier estimates of the cumulative risk of heart disease by time since diagnosis, in breast cancer patients and matched women from the general population. BC: breast cancer; REF: reference women in the matched controls from the general population; No.: number.

**Supplementary Table 1.** ICD codes used in the analyses.

| Disease codes | ICD-10 | ICD-9 |
| --- | --- | --- |
| Arrhythmia | I47, I48, I49 | 427 |
| Atrial fibrillation | I48 | 427D |
| Tachycardia and other cardiac arrhythmias | I47, I49 | 427A-C, 427E-J |
| Heart failure | I50 | 428A, 428B, 428X |
| Ischemic heart disease | I20-I25 | 410-414 |
| Myocardial infarction | I21,I22 | 410 |
| Angina pectoris | I20 | 411B, 413, 414A |
| Hypertension | I10-I15 | 401-405 |
| Tobacco abuse and Chronic Pulmonary Disease | J40-J47,J60-J67,F17,<br>I278,I279,J684,J701,J703 | 490-496,500-505,416W,416X,<br>506E,508B,508W,305B |

Abbreviations: ICD = International Classification of Diseases.

**Supplementary Table 2.** Cumulative incidence estimates of heart diseases in breast cancer patients and matched controls.

|  | Cumulative incidence, % (95% CI) |  |  |  |  |  |
| --- | --- | --- | --- | --- | --- | --- |
|  | 6 month | 1 year | 2 years | 5 years | 10 years | 15 years |
| Arrhythmia |  |  |  |  |  |  |
| Breast cancer | 0.86 (0.68-1.08) | 1.32 (1.10-1.59) | 1.92 (1.65-2.25) | 3.60 (3.21-4.04) | 7.10 (6.52-7.73) | 12.10 (11.12-13.16) |
| Matched control | 0.33 (0.29-0.37) | 0.60 (0.55-0.65) | 1.15 (1.08-1.23) | 2.78 (2.67-2.90) | 5.91 (5.75-6.08) | 9.60 (9.36-9.86) |
| Heart failure |  |  |  |  |  |  |
| Breast cancer | 0.20 (0.13-0.33) | 0.45 (0.33-0.62) | 0.75 (0.58-0.96) | 1.46 (1.21-1.75) | 2.93 (2.56-3.35) | 5.59 (4.91-6.37) |
| Matched control | 0.08 (0.06-0.10) | 0.17 (0.14-0.20) | 0.32 (0.29-0.36) | 0.86 (0.80-0.93) | 2.19 (2.09-2.29) | 4.37 (4.19-4.56) |
| Ischemic heart disease |  |  |  |  |  |  |
| Breast cancer | 0.46 (0.33-0.63) | 0.79 (0.62-1.00) | 1.30 (1.08-1.57) | 2.57 (2.24-2.95) | 4.43 (3.97-4.93) | 6.55 (5.85-7.34) |
| Matched control | 0.23 (0.20-0.27) | 0.54 (0.49-0.59) | 1.07 (1.00-1.14) | 2.45 (2.35-2.56) | 4.68 (4.54-4.83) | 7.01 (6.80-7.22) |

Abbreviations: CI = confidence interval. Cumulative incidence estimates of heart diseases in all breast cancer patients (N = 8338) at different time-points following the index date (= date of diagnosis in breast cancer patients). Cumulative incidence estimates are obtained from Kaplan-Meier survival analyses.

**Supplementary Table 3.** Hazard ratios for heart diseases by different adjuvant therapies and time since diagnosis

| Treatment variables | HR (95% CI) for Arrhythmia |  | HR (95% CI) for heart failure |  | HR (95% CI) for ischemic heart disease |  |
| --- | --- | --- | --- | --- | --- | --- |
|  | <10y | >10y | <10y | >10y | <10y | >10y |
| Radiotherapy |  |  |  |  |  |  |
| Right-sided | REF (1.00) | REF (1.00) | REF (1.00) | REF (1.00) | REF (1.00) | REF (1.00) |
| Left-sided | 0.94 (0.76-1.16) | 0.83 (0.59-1.16) | 1.02 (0.73-1.43) | 0.92 (0.57-1.48) | 1.10 (0.84-1.44) | 1.32 (0.74-2.36) |
| Missing |  |  |  |  |  |  |
| Chemotherapy |  |  |  |  |  |  |
| No | REF (1.00) | REF (1.00) | REF (1.00) | REF (1.00) | REF (1.00) | REF (1.00) |
| Anthracyclines-based | 1.02 (0.76-1.36) | 1.00 (0.58-1.73) | **1.84 (1.19-2.85)** | **2.31 (1.21-4.43)** | 1.09 (0.75-1.60) | 1.18 (0.48-2.90) |
| Anthracyclines +Taxanes | 0.75 (0.37-1.50) | 0.75 (0.10-5.62) | **5.08 (2.56-10.06)** | 0.00 (0.00-.) | 1.28 (0.57-2.86) | 0.00 (0.00-0.00) |
| CMF | 1.74 (0.93-3.25) | 0.72 (0.17-3.00) | 1.33 (0.47-3.73) | 0.69 (0.09-5.17) | 0.67 (0.24-1.85) | 0.92 (0.12-7.10) |
| Type unknown | 1.18 (0.86-1.63) | 1.09 (0.63-1.89) | **1.75 (1.06-2.86)** | 1.38 (0.66-2.89) | 1.47 (0.99-2.19) | 1.31 (0.52-3.27) |
| Missing |  |  |  |  |  |  |
| Hormone therapy |  |  |  |  |  |  |
| No | REF (1.00) | REF (1.00) | REF (1.00) | REF (1.00) | REF (1.00) | REF (1.00) |
| Tamoxifen | 1.02 (0.77-1.34) | 1.10 (0.69-1.77) | 0.76 (0.50-1.14) | 1.06 (0.58-1.96) | 0.82 (0.58-1.16) | 1.36 (0.60-3.09) |
| Aromatase inhibitors | 1.09 (0.81-1.48) | 1.05 (0.56-1.96) | 0.92 (0.60-1.41) | 1.62 (0.76-3.46) | 1.36 (0.93-1.97) | 1.39 (0.48-4.06) |
| Type unknown | 0.76 (0.49-1.20) | 1.19 (0.68-2.09) | 0.91 (0.49-1.69) | 1.08 (0.51-2.29) | 1.20 (0.75-1.92) | 1.44 (0.54-3.83) |
| Missing |  |  |  |  |  |  |
| Trastuzumab * |  |  |  |  |  |  |
| No | REF (1.00) | REF (1.00) | REF (1.00) | REF (1.00) | REF (1.00) | REF (1.00) |
| Yes | 1.48 (0.90-2.43) | 0.00 (0.00-0.00) | **2.26 (1.20-4.27)** | 0.00 (0.00-0.00) | 1.41 (0.76-2.59) | 4.43 (0.39-49.76) |
| Missing |  |  |  |  |  |  |

Abbreviations: Total No. refers to the total number of patients. N events refers to the number of observed cases. HR = hazard ratio; CI = confidence interval. Hazard ratios are estimated from Cox proportional hazards models with time since diagnosis as underlying time scale. Hazard ratios for model1 are adjusted for age and calendar period. Hazard ratios for model2 are multivariable adjusted including age at diagnosis (years), year of diagnosis (2001/2002, 2003/2004, 2005/2006, 2007/2008), menopausal status (premenopausal, postmenopausal, unknown), Charlson comorbidity index (0, 1, $\geq 2$ ), clinical stage (I, II, III, unknown), type of surgery (no, partial mastectomy, total mastectomy, unknown), history of the specific heart disease, hypertension, chronic pulmonary disease and tobacco abuse, and all treatment variables listed in the table. \* Treatment-specific analysis of trastuzumab was restricted to patients diagnosed between 2005 and 2008.

**Supplementary Table 4.** Hazard ratios for heart diseases by different adjuvant therapies, excluding patients with a history of heart diseases

| Treatment variables | Arrhythmia<br>(N = 570) |  | Heart failure<br>(N =243) |  | Ischemic heart disease<br>(N =307) |  |
| --- | --- | --- | --- | --- | --- | --- |
|  | N events | HR (95% CI) | N events | HR (95% CI) | N events | HR (95%) |
| Radiotherapy |  |  |  |  |  |  |
| Right-sided | 218 | REF (1.00) | 83 | REF (1.00) | 99 | REF (1.00) |
| Left-sided | 208 | 0.91 (0.75-1.10) | 87 | 1.01 (0.74-1.36) | 118 | 1.14 (0.87-1.49) |
| Missing | 10 |  | 6 |  | 5 |  |
| Chemotherapy |  |  |  |  |  |  |
| No | 376 | REF (1.00) | 133 | REF (1.00) | 207 | REF (1.00) |
| Anthracyclines-based | 83 | 1.04 (0.79-1.38) | 51 | **1.81 (1.22-2.70)** | 44 | 1.15 (0.78-1.68) |
| Anthracyclines +Taxanes | 10 | 0.92 (0.47-1.79) | 13 | **4.27 (2.18-8.35)** | 6 | 1.23 (0.51-2.94) |
| CMF | 6 | 0.85 (0.37-1.95) | 3 | 0.98 (0.30-3.17) | 2 | 0.51 (0.13-2.11) |
| Type unknown | 87 | 1.20 (0.89-1.61) | 38 | 1.42 (0.91-2.21) | 47 | 1.41 (0.95-2.10) |
| Missing | 8 |  | 5 |  | 1 |  |
| Hormone therapy |  |  |  |  |  |  |
| No | 89 | REF (1.00) | 49 | REF (1.00) | 43 | REF (1.00) |
| Tamoxifen | 298 | 0.99 (0.76-1.27) | 99 | 0.78 (0.54-1.12) | 142 | 0.98 (0.68-1.42) |
| Aromatase inhibitors | 119 | 1.06 (0.79-1.42) | 61 | 1.03 (0.68-1.55) | 85 | **1.48 (1.00-2.20)** |
| Type unknown | 56 | 0.91 (0.64-1.30) | 29 | 0.91 (0.55-1.50) | 36 | 1.17 (0.73-1.88) |
| Missing | 8 |  | 5 |  | 1 |  |
| Trastuzumab * |  |  |  |  |  |  |
| No | 129 | REF (1.00) | 39 | REF (1.00) | 60 | REF (1.00) |
| Yes | 18 | 1.21 (0.71-2.04) | 13 | **2.00 (1.02-3.92)** | 13 | 1.57 (0.82-2.99) |
| Missing | 93 |  | 39 |  | 52 |  |

Abbreviations: Total No. refers to the total number of patients. N events refers to the number of observed cases. HR = hazard ratio; CI = confidence interval. Hazard ratios are estimated from Cox proportional hazards models with time since diagnosis as underlying time scale. Hazard ratios are multivariable adjusted including age at diagnosis (years), year of diagnosis (2001/2002, 2003/2004, 2005/2006, 2007/2008), menopausal status (premenopausal, postmenopausal, unknown), Charlson comorbidity index (0, 1, $\geq 2$ ), clinical stage (I, II, III, unknown), type of surgery (no, partial mastectomy, total mastectomy, unknown), and all treatment variables listed in the table. \* Treatment-specific analysis of trastuzumab was restricted to patients diagnosed between 2005 and 2008.

**Supplementary Figure 1.**
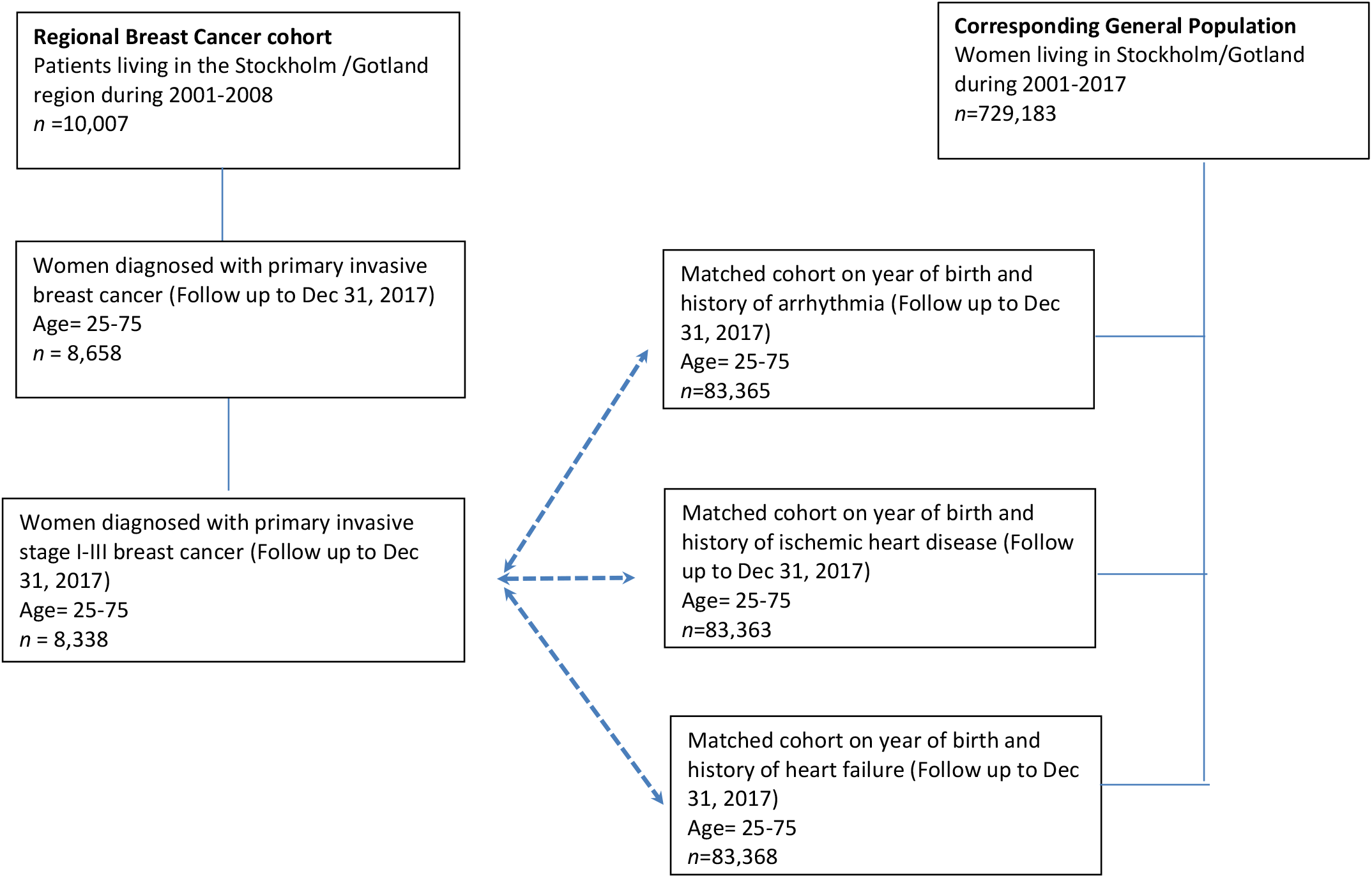
Flow chart of the data

**Supplementary Figure 2.**
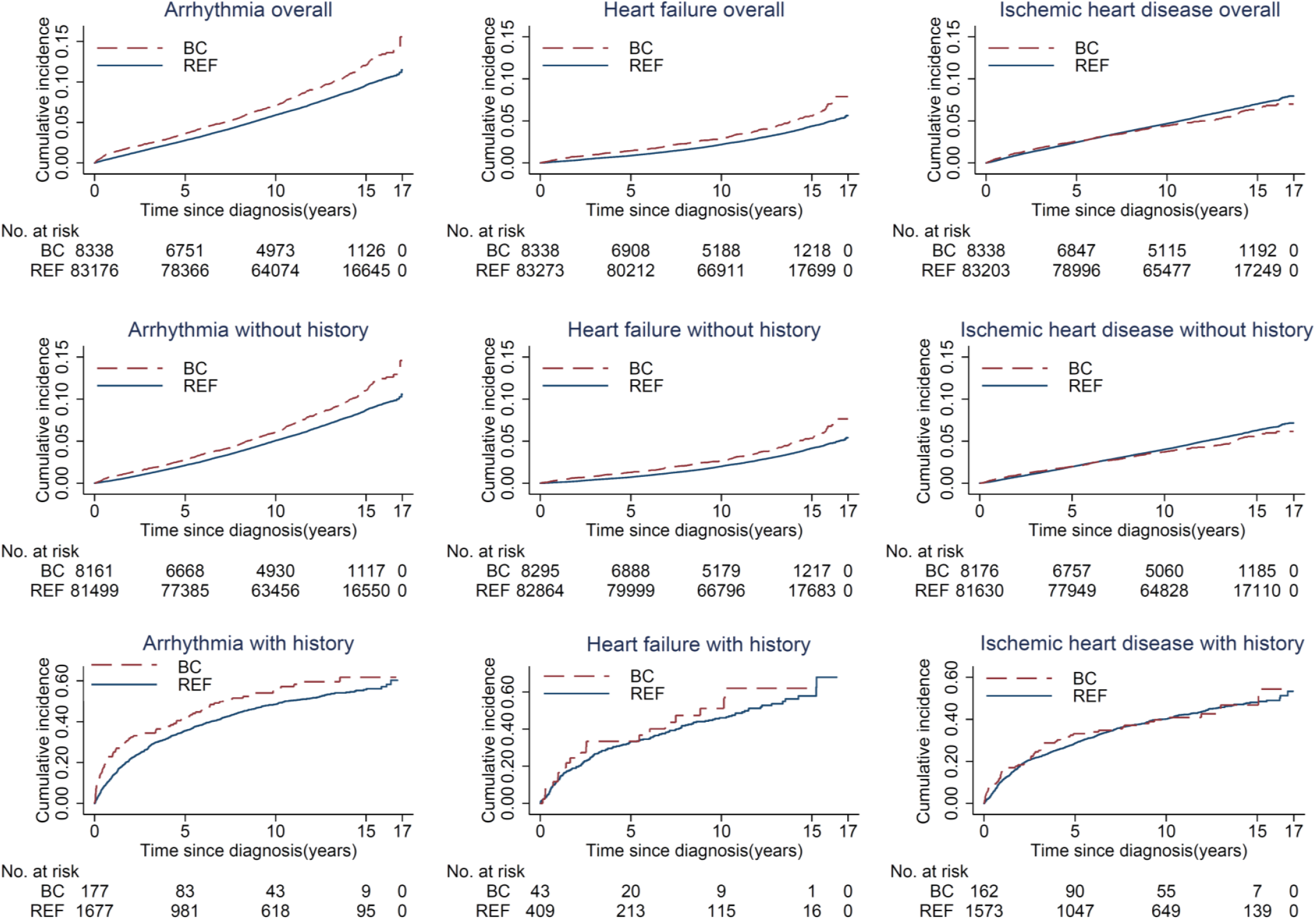
Cumulative incidence of heart diseases in breast cancer patients and matched reference individuals from the general population.

